# Exposure to unhealthy commodity brands in YouTube highlights of English Premier League and FIFA World Cup football matches

**DOI:** 10.64898/2026.08.17.26360621

**Authors:** Nathan Davies, Sarah Busby, Joanne Morling

## Abstract

**Background:** YouTube highlights packages are a major and growing route to football consumption, particularly among children and young people, but brand exposure within them has not been quantified. We measured unhealthy commodity brand exposure in English Premier League (EPL) and FIFA World Cup (WC) highlights.

**Methods:** We coded brand appearances lasting two or more seconds in 10 Sky Sports EPL highlights (final 10 games of the 2025/26 season) and 19 official FIFA 2026 WC highlights, recording commodity category, placement, and match moment, alongside pre-roll YouTube adverts. Data were collected between 4 June and 27 July 2026. Five highlights were double-coded (Cohen’s kappa 0.85).

**Results:** Overall brand density was similar across competitions (13.1 vs 13.9 references per minute), but composition differed markedly. Unhealthy commodity branding occupied 38.0% of EPL screen time versus 18.7% at the WC, a difference driven almost entirely by gambling (32.6% vs 1.5%). Gambling appeared in every EPL package, mainly on pitchside boards and LED screens (50.4%), with front-of-shirt accounting for 27.1%. WC exposure was more evenly spread across HFSS food (13%), alcohol (4%) and trading/crypto/prediction markets (3.7%), and appeared almost exclusively pitchside. Gambling brands accounted for ten of twelve pre-roll EPL adverts (123 of 153 seconds); no gambling adverts preceded WC highlights.

**Conclusions:** Gambling dominates unhealthy commodity exposure in EPL highlights, both in-video and in pre-roll advertising. Because most appearances occur away from the front of shirt, the voluntary front-of-shirt sponsorship withdrawal will leave the majority of this exposure intact. The WC comparison shows that tighter central control of the advertising environment produces lower and more diffuse exposure, and that governments and governing bodies with such control could restrict unhealthy categories altogether.

## BACKGROUND

The English Premier League (EPL) and the Fédération Internationale de Football Association (FIFA) World Cup (WC) represent two of the most-watched competitions for the world’s most popular sport, football. The EPL states that 1.87 billion people interact with Premier League content at least weekly through media.[1] The 2022 WC in Qatar was estimated to have engaged around 5 billion people across all media platforms, with 1.42bn people watching the final for at least one minute.[2] It is well-established that exposure to unhealthy commodities through advertising, including alcohol[3] and high fat, salt or sugar foods (HFSS)[4], is associated with increased consumption of these products. There is also growing evidence that sports-related gambling advertising is associated with increased betting frequency, expenditure and unplanned spend.[5,6]

Previous public health research has quantified alcohol and HFSS advertising exposure in samples of full matches during the 2018 WC, finding around 22% of programming time involved brand appearances for these products.[7,8] Gambling advertising was relatively rare.[8] In contrast, gambling advertising is highly prevalent in EPL broadcasts, where it is embedded through shirt sponsorship and hoardings rather than confined to commercial breaks. Frequency analyses of televised English professional football have identified 2.8 gambling references per broadcast minute, the large majority appearing in-play.[9] More recent research extended analysis to gambling-like products (in-play only) with gambling, cryptocurrency and financial-trading-app brands appearing 16, 7 and 3 times per broadcast minute respectively [10]

YouTube football highlights, typically 2 - 8 minutes long, have been an increasingly important channel for football consumption, especially for children and young people. 81% of 3-17-year-olds regularly watch YouTube.[11] UK residents can access Sky Sports’ free-to-view football highlights of every EPL game on YouTube as well as highlights of every WC game. Highlights condense the game down to incidents (such as goals and red cards) that are also most replayed on other media platforms (such as TikTok, Instagram, and traditional news media) and thus provide a convenient way to measure brand exposure for the most-watched moments. We present the first analysis of unhealthy commodity exposure for football YouTube highlights.

## METHODS

We measured brand exposure in (1) the Sky Sports YouTube highlights of the last 10 games of the 25/26 PL season and (2) one FIFA 2026 WC YouTube highlight from the official FIFA channel for each day of the first group stage match for each group, and knockout matches from the round of 16 onwards; if there were multiple games on a day, we selected the game featuring the highest-ranked team. Data collection took place between 4 June and 27 July 2026.

We coded the start and end time of a brand appearance for two or more seconds in the highlights, excluding kit manufacturer logos on kit (e.g. Adidas). We recorded its commodity category (alcohol, gambling/betting, HFSS food, sugary drink, energy drink, trading/crypto/prediction market). The Other category contained all other branding (e.g. automobiles, banking, hygiene products), For unhealthy commodities, we recorded its commodity subtype (e.g. regular, no/low alcohol, no/low sugar, alibi/brand extension, or “responsible consumption” message). We recorded brand placement, whether it was in the foreground or background, and the match moment (e.g. open play, goal).

Multiple brands could be recorded in the same match second, and if a brand disappeared for two or more seconds, then reappeared, this was coded as a new instance. Moving from one match moment to another (e.g. firstly, a passage of play leading to a goal, and secondly, a goal replay) initiated a new line for a brand.

We also coded the pre-highlight adverts that were served on YouTube before the highlights package was shown. A sample of 5 highlights were double-coded for interrater reliability, which was calculated on brand appearance using Cohen’s kappa. Disagreements were resolved by footage review by the first reviewer.

## RESULTS

Ten PL highlights and 17 WC top-ranked team highlights were coded. The EPL highlights had a mean length of 190 seconds; all but one of the WC highlights were 130 seconds long, with the highlights of the final 117 seconds long. The WC matches had approximately ten times the views of PL matches (a mean of 4.7 million compared to a mean of 450,000). Unhealthy commodity branding was visible during 38% of PL highlight duration compared to 18.7% of WC matches (Table 1).

**Table 1:** Summary of visible brands in Premier League and World Cup YouTube highlights.

| Measure | PL mean (SD) | WC mean (SD) |
| --- | --- | --- |
| Highlights coded (n) | 10 | 19 |
| Video duration (s) | 190.4 (11.4) | 129.1 (3.1) |
| Views (n) | 450,545 (287,810) | 4,777,036 (3,573,735) |
| Brand references per video (n) | 41.7 (13.0) | 29.9 (7.9) |
| Brand references per minute (n) | 13.1 (3.6) | 13.9 (3.6) |
| Distinct brands per video (n) | 15.2 (2.9) | 14.9 (3.9) |
| Unhealthy commodity appearances per video (n) | 17.4 (10.6) | 9.2 (4.1) |
| Unhealthy commodities appearances per minute (n) | 5.5 (3.2) | 4.3 (1.9) |
| Unique unhealthy brands per video (n) | 5.2 (1.6) | 5.5 (2.2) |
| Unhealthy commodity screen time (%) | 38.0 (21.2) | 18.7 (9.1) |

**Table 2:** Presence of commodity category by competition.

| Competition | Commodity category | Presence in a highlights package (n) | Unique appearances (n) | Time present on screen (%) |
| --- | --- | --- | --- | --- |
| Premier League | Alcohol | 5 | 6 | 1.7 |
|  | Energy drink | 6 | 15 | 3.3 |
|  | Gambling/betting | 10 | 129 | 32.6 |
|  | HFSS food | 2 | 7 | 2.5 |
|  | Other | 10 | 243 | 38.7 |
|  | Trading/crypto/prediction market | 3 | 17 | 4.4 |
| World Cup | Alcohol | 11 | 25 | 4 |
|  | Energy drink | 0 | 0 | 0 |
|  | Gambling/betting | 7 | 10 | 1.5 |
|  | HFSS food | 17 | 93 | 13 |
|  | Other | 17 | 350 | 44.1 |
|  | Trading/crypto/prediction market | 15 | 29 | 3.7 |

**Table 3:** Proportional location of unhealthy commodity branding by competition.

| Competition | Commodity category | Pitchside board/LED | Fan | Front of shirt | Player or staff equipment | Sleeve | Stadium signage | Manager/staff |
| --- | --- | --- | --- | --- | --- | --- | --- | --- |
| Premier League | Alcohol | 83.3 | 16.7 | 0 | 0 | 0 | 0 | 0 |
|  | Energy drink | 66.7 | 0 | 20 | 6.7 | 6.7 | 0 | 0 |
|  | Gambling/betting | 50.4 | 3.1 | 27.1 | 0.8 | 5.4 | 10.1 | 3.1 |
|  | HFSS food | 85.7 | 0 | 0 | 0 | 0 | 14.3 | 0 |
|  | Trading/crypto/prediction market | 64.7 | 0 | 0 | 0 | 35.3 | 0 | 0 |
| World Cup | Alcohol | 100 | 0 | 0 | 0 | 0 | 0 | 0 |
|  | Gambling/betting | 100 | 0 | 0 | 0 | 0 | 0 | 0 |
|  | HFSS food | 97.8 | 0 | 0 | 2.2 | 0 | 0 | 0 |
|  | Trading/crypto/prediction market | 100 | 0 | 0 | 0 | 0 | 0 | 0 |

**Table 4:**
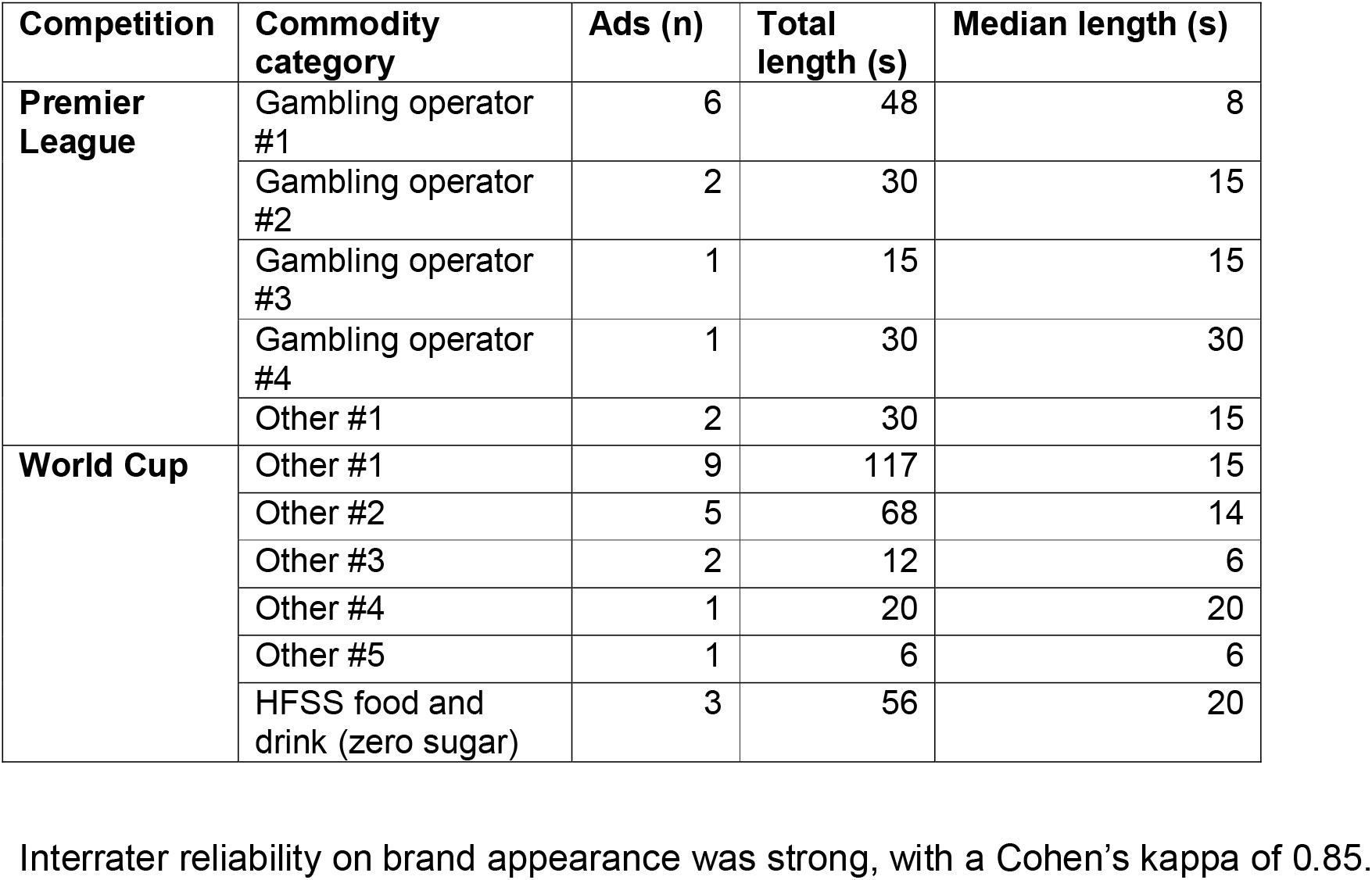
YouTube adverts preceding highlights.

| Competition | Commodity category | Ads (n) | Total length (s) | Median length (s) |
| --- | --- | --- | --- | --- |
| **Premier League** | Gambling operator #1 | 6 | 48 | 8 |
|  | Gambling operator #2 | 2 | 30 | 15 |
|  | Gambling operator #3 | 1 | 15 | 15 |
|  | Gambling operator #4 | 1 | 30 | 30 |
|  | Other #1 | 2 | 30 | 15 |
| **World Cup** | Other #1 | 9 | 117 | 15 |
|  | Other #2 | 5 | 68 | 14 |
|  | Other #3 | 2 | 12 | 6 |
|  | Other #4 | 1 | 20 | 20 |
|  | Other #5 | 1 | 6 | 6 |
|  | HFSS food and drink (zero sugar) | 3 | 56 | 20 |

Interrater reliability on brand appearance was strong, with a Cohen's kappa of 0.85.

## DISCUSSION

This is the first analysis of unhealthy commodity brand exposure in football YouTube highlights. Overall brand density was similar across the two competitions at around 13 references per broadcast minute, but the composition of that exposure differed markedly. Unhealthy commodity branding occupied 38.0% of EPL screen time compared with 18.7% for the WC, a difference driven almost entirely by gambling advertising, which accounted for 32.4% of PL screen time and 1.5% of WC screen time. The WC figure is close to the approximately 22% of programming time attributed to alcohol and HFSS branding in full-match broadcasts of the 2018 tournament.[7,8]

The format of highlights packages itself influences the brands displayed. Highlights compress a match into goals, near-misses and disciplinary incidents, rather than close-ups of players or managers during lull times in matches. This means they disproportionately retain passages of goalmouth and replay footage in which perimeter boards and LED hoardings fill the frame. Pitchside boards and LED screens carried half of all PL gambling appearances and all WC gambling, alcohol, HFSS and trading appearances. Given that these are also the moments most redistributed across platforms such as TikTok and Instagram, and traditional news media, this holds significance for total population exposure.

Gambling exposure in the EPL extended beyond the highlights themselves. Researchers encountered up to 30 seconds of dedicated gambling advertising before roughly 190 seconds of content, meaning pre-roll advertising alone approached a fifth of the total viewing episode. Our findings add to the evidence that the voluntary withdrawal of gambling sponsorship from the front of EPL shirts is insufficient to tackling gambling harms. Front-of-shirt placement accounted for 27.1% of gambling appearances, while pitchside boards, stadium signage, sleeves and staff clothing together accounted for the remaining 72.9%. Removing front-of-shirt branding would therefore leave most of the exposure in these highlights untouched, and the continued availability of sleeve sponsorship and advertising hoardings offers an obvious substitution route. Measures confined to a single placement are unlikely to produce meaningful reductions in gambling advertising exposure.

The WC comparison demonstrates that a different distribution is achievable. FIFA exercises close control over stadium advertising, requiring venues to obscure brands that are not tournament partners, and the resulting brand appearance is both smaller and more evenly spread across categories. However, an organisation able to eliminate non-partner branding entirely is equally able to decide which categories appear at all. The significant presence of alcohol, HFSS food, sugary drinks and cryptocurrency and prediction market sponsors is a FIFA decision. FIFA – or in the case of the EPL, the UK government - could exercise control over the advertising environment to entirely restrict brand appearances to neutral or health-promoting categories.

## Limitations

This was a purposive sample of 27 highlights packages and cannot be taken as representative of either competition across a full season or tournament. Views data indicate audience size but not composition, so we cannot estimate the proportion of child viewers. Our two-second threshold for a codeable appearance is conservative and will have excluded briefer exposures, particularly rapid camera cuts during replays; appearance rates are consequently not comparable with studies counting every simultaneous logo appearance. Pre-roll advertising on YouTube is personalised and served on the basis of individual user profiles; this study reports one experience.

## Conclusions

Gambling occupied 32.4% of EPL highlight screen time, appeared in every package, and added 123 seconds of pre-roll advertising, against 1.5% and none at the World Cup. With 50.4% of EPL gambling appearances on pitchside boards and LEDs and only 27.1% front of shirt, the voluntary front-of-shirt withdrawal will leave most of this exposure intact.

## Ethics statement

No ethical approval was required for this analysis of publicly available broadcast content.

## Data availability statement

All data produced in the present study are available upon reasonable request to the authors.

